# The uncemented Avenir^®^ stem covered with hydroxyapatite in a septic environment in the revision of an infected total hip arthroplasty : a report on 40 cases

**DOI:** 10.1101/2022.09.05.22279579

**Authors:** Matthieu Mangin, Zouhair Aouzal, Grégoire Leclerc, Anne Pauline Sergent, Kévin Bouiller, Isabelle Patry, Patrick Garbuio

## Abstract

Does the implantation of an uncemented hydroxyapatite-coated first-line stem in a septic environment during a one-stage total hip arthroplasty revision (THAR) for periprosthetic joint infection on total hip arthroplasty provide good results in terms of healing the infection and osteointegration of the stem ?

We retrospectively reviewed 40 patients operated on for septic THAR with placement of the cementless Avenir® stem - between 2008 and 2018 at the Besançon University Hospital - with a minimum follow-up of 2 years necessary to define cure in the absence of infectious recurrence. Clinical outcome was assessed using the Harris, Oxford and Merle D’Aubigné scores. Osteointegration was analyzed by the Enhg radiographic score.

Mean follow-up was 4.5 years (0-11). Cure of infection was achieved in 35 of 40 (87.5%). The median Harris score was 74/100, Oxford score 45/60, and Merle d’Aubigné score 15/18. Of 37 femoral stems, 36 (97%) had radiographically stable osteointegration. An age of 80 years is a risk factor for failure of septic THAR with implantation of an uncemented stem in one stage. The cementless Avenir® stem has its place in one-stage septic THAR. It gives good results on the healing of the infection and the integration of the stem in the context of femoral bone loss rated Paprosky 1.

## 1. Introduction

Faced with a periprosthetic joint infection on total hip arthroplasty the surgeon may decide to change the whole prosthesis with a two-stage surgery which has long been the reference (1).

Currently, the one-stage technique ensures a rate of healing of the infection identical to the two-stage technique, in the order of 90% (2), ensures better functional results (3,4) and a better quality of life (5).

The published studies on 1-stage septic THAR mainly deal with the implantation of a cemented stem with antibiotic-impregnated cement (6). Some authors even advise against the use of a cementless stem (7).

Some manufacturers advise against using their uncemented stem in septic environments. The main objective of this study is to determine whether the implantation of an uncemented hydroxyapatite-coated first-line stem in a septic setting during a one-stage infected THAR procedure has a good outcome on the healing of the infection.

The secondary objectives are to evaluate the integration of the pressfit stem in infected and remodeled bone, the functional and satisfaction scores of the operated patients and to identify potential risk factors for failure of this surgical technique.

## 2. Material and method

We conducted a retrospective observational monocentric study in the unit of the Reference Center for Complex Osteoarticular Infections at the Besançon University Hospital from 2008 to 2018.

Inclusion criteria were: a periprosthetic joint infection to the hip according to the Musculoskeletal Infection Society (MSIS) criteria (8), femoral bone loss Paprosky 1 (9), septic THAR with bipolar change and implantation of a cementless first-line stem covered with hydroxyapatite, according to the one-stage technique or the two-stage technique if the intraoperative bacteriological samples were still positive at the second stage.

The exclusion criteria were: septic THAR using the 2-stage technique with sterile bacteriological sampling in the second stage, the use of revision or reconstruction or cemented stems, and intermediate hip arthroplasty.

In these THAR, we implanted the cementless Avenir® Uncemented stem (Zimmer Biomet, Warsaw, USA), which was completely covered with hydroxyapatite. The acetabulum was cemented or uncemented according to the surgeon’s intraoperative findings.

In our management, the total duration of antibiotic therapy was 3 months. We initiated suppressive antibiotic therapy in 5 of our patients.

The minimum follow-up time for all patients, excluding failures, was 2 years.

The primary endpoint was successful treatment of the infection, defined as no recurrence of infection during the minimum two years of follow-up after revision surgery, with clean scarring and a CRP < 5mg/l in a patient who could be on suppressive antibiotic therapy if this had been decided initially at the Multidisciplinary Consultation Meeting on Osteoarticular Infections for prophylaxis.

The secondary endpoints were radiological integration of the prosthesis according to the Engh score (10,11), evaluation of hip function scores (Harris, Oxford, Merle d’Aubigné modified by Postel (PMA) and Charnley), SF12 quality of life score, patient satisfaction score on a scale of 0 to 10, Patient Acceptable Symptom State (PASS) (12) and Minimal Clinically Important Difference (MCID) (13).

We used the software Excel® and BiostaTGV for data collection and statistical analysis. In order to search for risk factors of failure of our technique, we have performed Fischer tests (N < 5) for qualitative variables (95% CI) and Student tests (N < 30) for quantitative variables (p < 0.05).

## 3. Results

We included 40 patients whose characteristics are described in Table 1. 35 patients received a standard uncemented stem in one stage and 5 received a two-stage revision with positive intraoperative samples in the second stage, resulting in the implantation of the stem in a septic environment.

**Table 1:**
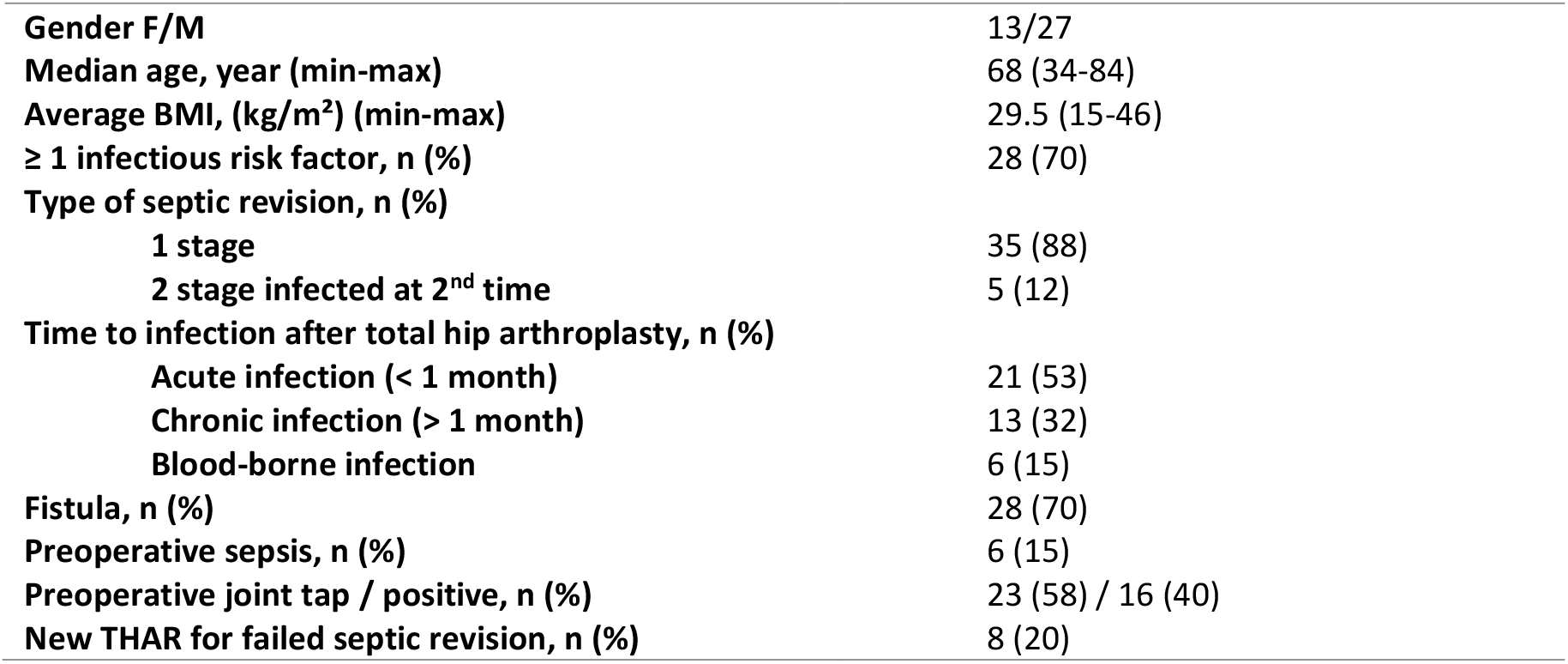
Characteristics of the 40 patients in the study.

Of the 40 THAR, 30 (75%) were positive for a single bacterium and the remaining 10 (25%) were positive for 2 or even 3 bacteria (Table 2).

**Table 2:** Microbiological analysis of intraoperative samples of septic THAR.

| Bacteria | N (%) |
| --- | --- |
| **Staphylococcus** | 32 (59) including 8 resistant |
| meticillin-sensitive *aureus* | 9 (17) |
| meticillin-resistant *aureus* (MRSA) | 2 (4) |
| meticillin-sensitive *epidermidis* | 10 (18) |
| meticillin-resistant *epidermidis* (MRSE) | 6 (11) |
| Other | 5 (9) |
| **Enterococcus faecalis** | 4 (7) |
| **Enterobacteria** | 8 (15) |
| *Enterobacter cloacae* | 5 (9) including 1 ESBL |
| *Klebsiella pneumoniae* | 1 (2) |
| *Escherichia coli* | 2 (4) |
| **Pseudomonas aeruginosa** | 3 (6) |
| **Other** | 7 (13) including 1 resistant |
| **Total** | 54 |

Of the 40 patients, 35 (87.5%) showed healing of their periprosthetic joint infection without recurrence at a minimum of 2 years follow-up and a mean of 4.5 years (0-11).

Among the 5 failures, there were 4 cases of persistent infections between 0 and 4 months. Two patients underwent an enlarged synovectomy with washing and replacement of moving parts associated with suppressive antibiotic therapy, one of whom recovered and the other did not. The third patient refused to undergo surgery because of the persistence of a fistula and was put on suppressive antibiotic therapy which resulted in healing without infectious recurrence. The fourth patient underwent a new one-stage THAR with insertion of a new uncemented stem which resulted in healing of the infection. Finally, the last patient died 1 month after the THAR from a postoperative complication.

Radiographs were analyzed in 37 patients (35 successes and 2 failures currently cured by synovectomy lavage and/or suppressive antibiotic therapy). 36 patients (97%) showed good osteointegration of the stem. All acetabular cups were osseointegrated or cemented. Only one patient had a revision at four years from our THAR, for mechanical loosening of the stem with sterile bacteriological samples at the time of revision.

The analysis of the secondary endpoints was carried out on 28 patients (Table 3). Half of the patients were satisfied with our care with a satisfaction score of 9/10 or higher. 19 patients (68%) described a satisfactory current health status for the coming years at PASS and 23 patients (82%) felt that the treatment of their periprosthetic joint infection by our method improved their health status at MCID.

**Table 3:**
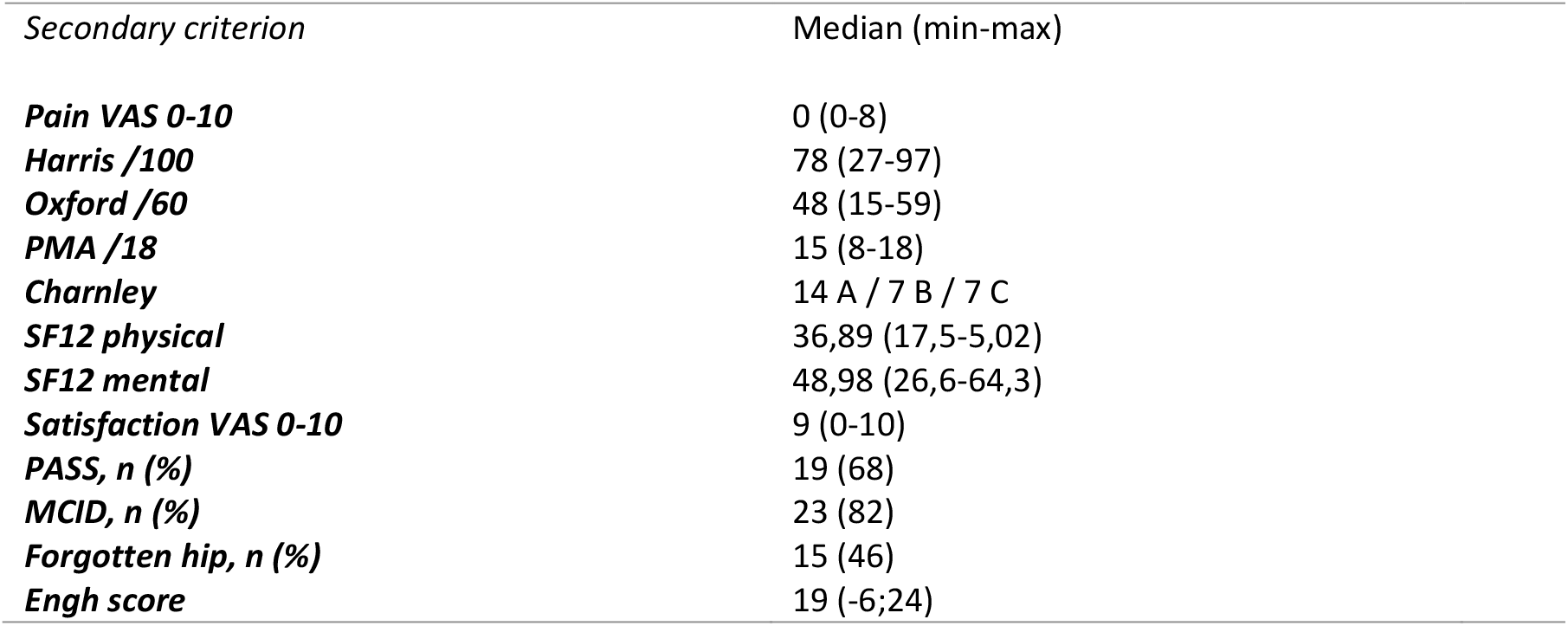
Secondary endpoint results for 28 patients.

Comparison between the failed and successful THAR groups shows that patients with failure are older at 80.6 years [78.58 - 82.62] versus 65.8 years [61.59 - 70.00] for success (p=0.014). The search for risk factors shows that only an age ≥ 80 years is a risk factor for failure of our THAR technique (Odds Ratio 26.56 [2.05 - 1553], p=0.003) (Table 4).

**Table 4:** Fisher’s exact test for risk factors for THAR failure.

| FACTOR STUDIED | ODDS RATIO; 95% CI | P-VALUE |
| --- | --- | --- |
| **AGE ≥ 80 YEARS** | **26,56 [2,05 - 1553]** | **0,00348** |
| CHRONIC INFECTION ≥ 1 MONTH | 1,76 [0,18 - 23,49] | 0,0654 |
| HISTORY OF FAILED SEPTIC REVISION | 1 [0,001 - 12,51] | 1 |
| NO IDENTIFICATION OF BACTERIA PREOPERATIVELY | 0,43 [0,03 - 4,02] | 0,3725 |
| BACTERIA RESISTANT TO INTRAOPERATIVE BACTERIOLOGY | 3,82 [0,26 - 43,55] | 0,2039 |
| PREOPERATIVE FISTULA | 0,69 [0,07 - 9,43] | 1 |
| PREOPERATIVE SEPSIS | 1,48 [0,03 - 20,05] | 1 |

## 4. Discussion

After a mean follow-up of 4.5 years, the infection cure rate is 87.5% with implantation of the hydroxyapatite-coated uncemented Avenir® stem in a septic setting. These results are identical or better than published studies on septic THAR. The literature reports a cure rate for this technique ranging from 56% to 95% (14–19) (Table 5). One-stage surgery with a cemented stem has a cure rate of 75-90% (2,20). Two-stage surgery, with or without a cemented stem, has a cure rate of 76-90% (4,21).

**Table 5:** Bibliography on 1-stage septic THAR with uncemented stem.

| 1st Author | Ref. | Year | Nbr. of patients | Average follow-up (years) | Type of study | Cure of infection (%) | Note |
| --- | --- | --- | --- | --- | --- | --- | --- |
| George | (14) | 2016 | 148 | 3 | Literature review | 86 | Healing identical to the 2-stage and 1-stage cemented |
| Yoo | (15) | 2009 | 12 | 7 | Retrospective | 83 |  |
| Hansen | (16) | 2013 | 27 | 4 | Retrospective | 56 | 70% cure including washings after THAR |
| Bori | (17) | 2013 | 42 | 4 | Retrospective | 95 |  |
| Ji | (18) | 2019 | 126 | 5 | Retrospective | 89 | Vancomycin in situ |
| Lange | (19) | 2017 | 56 | 4 | Prospective | 91 | Vancomycin + Gentamycin in situ |

The results of our study are all the more satisfactory since two of the five patients who failed acquired healing without new recurrence after synovectomy, lavage, change of moving parts and suppressive antibiotic therapy for one and suppressive antibiotic therapy only for the other.

The osteointegration rate of the hydroxyapatite stem in a septic environment at the last follow-up of 97% is identical to those found in the literature (18). The good integration of the stem supports the success of our surgical management.

Compared to the literature, the functional and satisfaction scores of the patients in the study are in the middle of the published literature (17,18,22,23).

If in our study, a patient aged 80 years and over has a statistically higher risk of septic THAR failure with infectious recurrence, there is no evidence that the 2-stage technique would give a better result in this group of patients.

Eight patients in our study had a history of failed septic THAR where we attempted a new septic revision. Two had a new failure without this being a significant risk factor. Caution should be exercised when performing a septic THAR in a patient with a previous failed septic revision. A 2-stage or 1-stage management with a cemented stem could then be discussed (20).

Six patients had sepsis on THAR. For MSIS (24), this is a relative contraindication to one-stage management. Two of them failed our management without being a significant risk factor. However, the management of a patient with sepsis or septic shock on a periprosthetic joint infection must be adapted. An evacuation puncture or even an emergency drainage with multiple bacteriological samples and the implementation of a broad spectrum antibiotic therapy should be discussed. Complete prosthetic replacement or replacement of the moving parts with extended synovectomy should only be performed at a later stage after control of the organ dysfunctions.

In our study, the presence of a fistula was not a risk factor for septic THAR failure. These data suggest that the presence of a fistula is not a contraindication to a one-stage change in septic THAR, contrary to what is sometimes recommended (24).

The absence of preoperative bacteriological identification was not a factor in the failure of our technique, especially since the presence of resistant bacteria in the preoperative bacteriology did not prevent the healing of the infected prosthesis. *The AAOS* published in 2010 (25) an algorithm for the diagnosis of prosthetic infections, advised by the MSIS (24), in which it is recommended to perform a preoperative puncture only in patients with painful hip symptoms with an inflammatory syndrome and in the absence of a fistula.

Of the 9 patients with resistant bacteria, 7 had successful septic THAR (2 failures with MRSE and multidrug-resistant *Corynebacterium*). Thus, we had a good cure rate for MRSA (2/2, 100%) and MRSE (5/6, 83%). Hischebeth et al. (26) report in 2019 a cure rate in 2-stage septic THAR of 80% for MRSA and only 54% for MRSE. The presence of resistant bacteria is not a risk factor for failure in our study. Contrary to the recommendations (20,24), preoperative isolation of resistant bacteria (puncture, previous surgery) is not necessarily a contraindication to the use of a 1-stage technique.

The limitation of our study is that it is retrospective with no control group. The small number of patients included in the study does not allow us to analyse all the possible risk factors for failure of our technique. However, in comparison with the literature, our study has a larger number of patients operated on with a longer follow-up and does not use local antibiotic therapy administered in the femoral shaft before implantation of the stem, which is done in several studies (18,19).

## 5. Conclusion

The uncemented, hydroxyapatite-coated Avenir® first-line stem has its place in one-stage septic THAR.

It gives good results in terms of healing of the infection and integration of the stem in the case of femoral bone loss rated Paprosky 1. The results obtained are identical to those obtained in one-stage management with a cemented prosthetic stem or in two-stage management, without the complications of the latter two techniques.

The results of this study showed that this surgical technique is possible, even in the presence of a fistula, a resistant germ or the absence of preoperative germ identification.

## Data Availability

All data produced in the present study are available upon reasonable request to the authors

